# Large Language Models for Individualized Psychoeducational Tools for Psychosis: A cross-sectional study

**DOI:** 10.1101/2024.07.26.24311075

**Authors:** Musa Yilanli, Ian McKay, Daniel I. Jackson, Emre Sezgin

## Abstract

**Importance:** In mental healthcare, the potential of Large Language Models (LLMs) to enhance psychoeducation is a burgeoning field. This study explored the potential of ChatGPT as an individualized psychoeducational support tool specifically for psychosis education.

**Objective:** The study aims to evaluate psychosis-related questions to provide accurate, clear, and clinically relevant individualized information for patients and caregivers.

**Design:** This cross-sectional study uses a qualitative analysis design. The researchers specifically employed a question-answering system (GPT-4 via ChatGPT) to generate responses to common questions about psychosis. Experts in the field then evaluated these responses to assess their quality for use in a clinical setting.

**Primary Outcome:** Researchers presented ChatGPT with 20 common questions frequently asked by patients’ caregivers and relatives. Two experts in psychosis then assessed the quality of the responses using six criteria: accuracy (1-3), clarity (1-3), inclusivity (1-3), completeness (0-1), clinical utility (1-5) and an overall score (1-4).

**Results:** The evaluation yielded positive results overall. Responses were rated as accurate (M±SD= 2.89±0.22) and clear (mean score of 2.93±0.18). There was potential for improvement in terms of inclusivity (mean score of 2.30±0.41), suggesting a need to incorporate more diverse perspectives. Completeness received high ratings (mean score of 0.93±0.18), indicating responses addressed all aspects of the questions. Most importantly, the responses were deemed clinically useful (mean score of 4.35±0.52).

**Conclusions:** In summary, this study underscores the significant promise of ChatGPT as a psychoeducational tool for patients with psychosis, their relatives, and their caregivers. The experts’ findings affirm that the information delivered by ChatGPT is not only accurate and clinically relevant but also conveyed conversationally, enhancing its accessibility and usability. The initial performance of ChatGPT as a psychoeducational tool in the context of psychosis education is undeniably positive.

## Introduction

Psychosis, characterized by a detachment from reality, presents a significant challenge for individuals and healthcare systems alike [1].Effective education empowers patients and their caregivers to understand their condition, manage symptoms, and navigate the complexities of treatment. Traditionally, healthcare providers served as the primary source of medical information for patients and caregivers. Individuals may also seek education through reliable sources or utilize search engines. However, readily accessing reliable information sheets for specific questions about symptoms, diagnosis, treatment, and outcomes can be a time-consuming task. Recently, AI chatbots have emerged as a potential solution, offering convenient and readily available health information. LLMs, trained on vast datasets of text and code, hold immense promise for developing interactive and adaptable learning tools [2, 3]. A crucial concern regarding LLM-generated information is its accuracy and reliability [3]. The quality of training data significantly influences LLM output, and the vast amount of online information presents challenges in ensuring factual accuracy [4, 5].

To address these concerns, this research investigated the LLM’s capability to answer frequently encountered clinical questions from real-world patients and caregivers pertaining to psychosis. The assessment centered on five critical aspects: factual accuracy, clarity of communication, language inclusivity, information comprehensiveness, and the overall clinical usefulness of the LLM’s responses. This evaluation offers valuable insights into the potential application of LLM-powered chatbots for providing tailored psychosis education.

## Methods

We used GPT-4 (via ChatGPT UI) as the LLM for this study. We curated 20 psychotic disorder questions based on anecdotal evidence, as frequently asked and documented in pediatric behavioral health clinics (See Appendix for the question list). Then, ChatGPT was prompted with each question in a new single session. We ensured response consistency by re-prompting the same question three times and comparing the responses (first three questions). One board-certified clinical psychiatrist and a clinical psychologist (Co-authors MY, and IM) assessed and rated the responses. We used a 6-category rating rubric including accuracy, clarity, inclusiveness, completeness, clinical utility, and overall score (Table 1). The Rubric categories were informed by the literature (See Table 1). Each generative response was measured for readability and demographic characteristics using the SHeLL Editor on default settings.

**Table 1.** Categories, scales, and descriptions of this rating scale.

| Rubric Category | Question | Description | Grading Scale |
| --- | --- | --- | --- |
| **Accuracy** | Is the answer an accurate reply to the question? | Assesses if the response correctly addresses the question. "Accurate" fully answers with correct info. "Inaccurate" is incorrect or irrelevant. "Partially accurate" is somewhat correct but incomplete. | Accurate, Partially Accurate, Inaccurate (1-3) |
| **Clarity** | Is the message clearly conveyed? | Measures how understandable the response is. "Yes" is well-structured and clear. "No" is confusing or complex. "Partially" is somewhat clear but needs improvement. | Yes, Partially, No (1-3) |
| **Inclusivity** | Is the message inclusive for a diverse range of recipients regardless of race/ethnicity, culture, etc.? | Evaluates cultural sensitivity and appropriateness. "Yes" respects diversity. "No" is specific to one group. "Partially" shows some effort but needs improvement. | Yes, Partially, No (1-3) |
| **Completeness** | Does the response completely answer the question? | Assesses if the response fully covers the question. "Complete" covers all aspects. "Incomplete" misses critical elements. | Complete, Incomplete (0-1) |
| **Clinical Utility** | Is the response practical in a clinical context? | Measures practical usefulness in clinical settings. Ranges from "Strongly Disagree" to "Strongly Agree." | Strongly Agree, Agree, Neutral, Disagree, Strongly Disagree (1-5) |
| **Overall Score** | What is the overall score? | Reflects confidence in accuracy, clarity, inclusivity, completeness, and utility. "Very Low" has major issues. "Low" has notable deficiencies. "Moderate" is reliable with minor improvements needed. "High" shows strong confidence. | High, Moderate, Low, Very Low (1-4) |

## Results

The evaluation of responses across different dimensions—accuracy, clarity, inclusivity, completeness, and clinical utility—reveals a nuanced picture of their quality and applicability (see Figure 1, graph illustrates normalized scores). The accuracy (mean = 2.88, SD = 0.22) and clarity (mean = 2.93, SD = 0.18) scores indicate that, on average, responses are mostly accurate and clear, suggesting they generally address the questions correctly and are communicated in an understandable manner. Low standard deviations indicate a consistency in scoring for these categories. However, inclusivity (mean = 2.30, SD = 0.41) presents a notable departure from these strengths, indicating a moderate consideration for a diverse audience. Completeness scores (mean = 0.93, SD = 0.18) indicate each response has been perceived to include comprehensive information. Clinical utility scores (mean = 4.35, SD = 0.52) are notably high, implying that the responses, despite their shortcomings in inclusivity, provide substantial practical value in a clinical context. The overall score (mean = 3.55, SD = 0.47) reflects an above moderate level of confidence in the responses’ quality and applicability across the assessed dimensions (Figure 1). The readability performance of this tool is consistent and falls within a reasonable range with the average readability score of 15.59 (SD=1.59, IQR= 1.9), indicating a generally moderate level of readability (Appendix 1).

**Figure 1.**
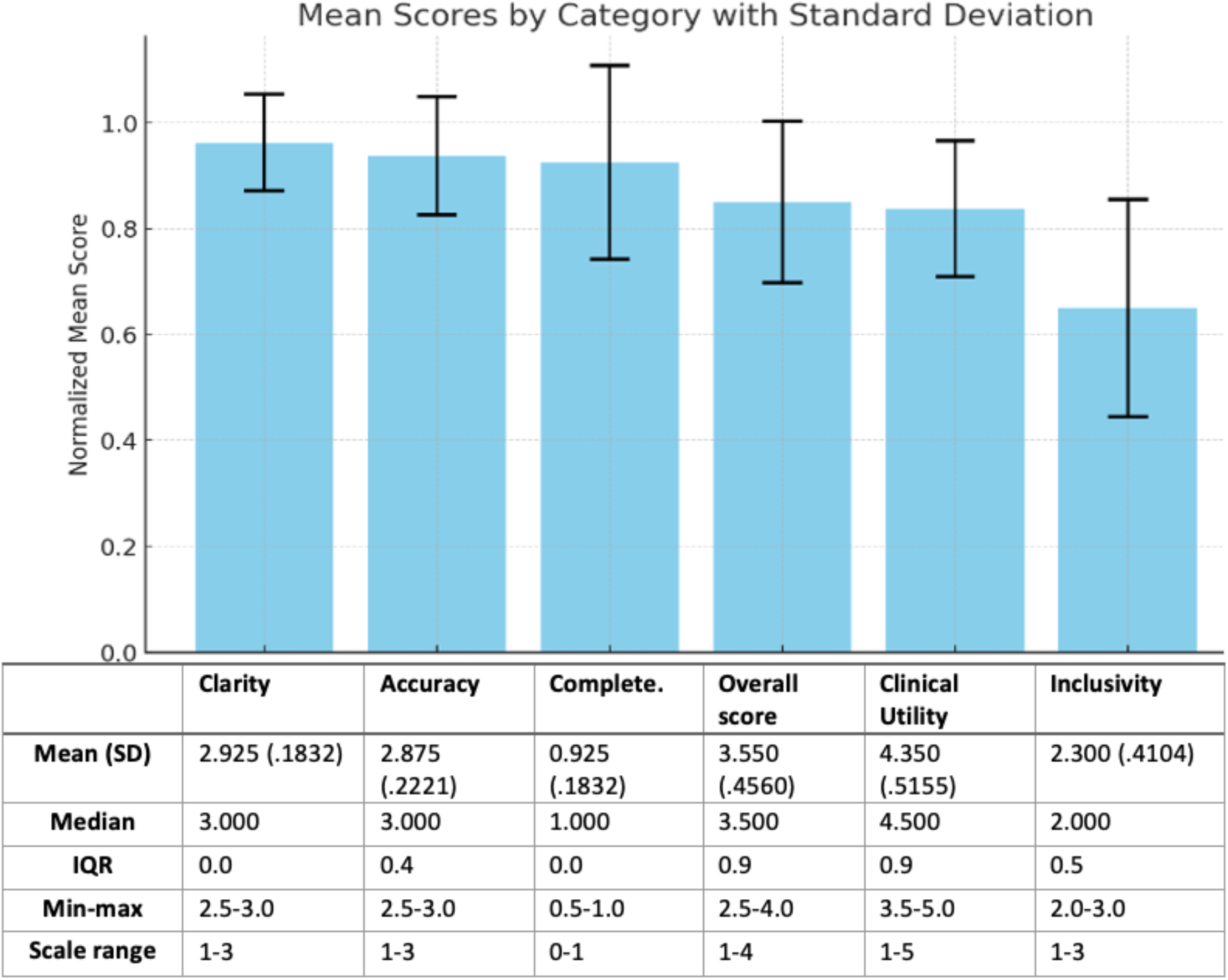
Mean scores by categories

## Discussion

The utilization of ChatGPT for the development of psychoeducational materials for Psychosis represents a novel approach to improving the accessibility and dissemination of information in clinical settings. Overall, this evaluation presents evidence that ChatGPT could serve as a supplement for psychoeducational tools for individuals with psychosis-related inquiries, including patients, relatives, and caregivers. The readability score indicates that comprehension of the content could be challenging for an average reader. Overall, the results are promising, with GPT-4 demonstrating high accuracy and clarity in answering common clinical questions, suggesting that ChatGPT4 effectively synthesized information to provide accurate and comprehensible responses to psychosis-related inquiries. These findings are consistent with previous research highlighting the capabilities of LLM in understanding and generating impactful responses to common inquiries about medical diagnoses [7]. However, the inclusivity of diverse perspectives needs improvement, underscoring the importance of considering a broader range of cultural, social, and experiential contexts in developing psychoeducational materials. Of note, the wording of our question prompts may have impacted the LLM’s sensitivity to include cultural factors in their responses Despite this limitation, responses were rated highly for completeness, indicating they answered all aspects of the question. Lastly, these responses were deemed clinically useful, highlighting the potential of clinical settings. Further studies are suggested to explore if LLM may offer a more accessible and less time-consuming experience compared to conventional resources and practices.

## Conclusion

Leveraging large language models (LLMs) like GPT-4 as a psychoeducational tool in psychosis holds promise for patients, relatives and caregivers. Our results highlight the promise of ChatGPT -based tools for delivering accurate and accessible educational resources for patients with psychosis. While traditional resources may struggle with these aspects, ChatGPT offers a potentially more approachable format. Continued research and development of AI technologies for mental health education are crucial to further optimize their effectiveness and ensure inclusivity.

## Data Availability

All data produced in the present study are available upon reasonable request to the authors

## Appendix Readability scores

Appendix: Readability scores
| # | readability | characters | words | unique words | sentences |
| --- | --- | --- | --- | --- | --- |
| 1 | 15.8 | 2427 | 326 | 158 | 31 |
| 2 | 18.1 | 2244 | 323 | 168 | 21 |
| 3 | 12.5 | 1778 | 230 | 152 | 31 |
| 4 | 16.3 | 2656 | 338 | 166 | 32 |
| 5 | 13.7 | 2949 | 427 | 233 | 38 |
| 6 | 14.7 | 3523 | 481 | 218 | 46 |
| 7 | 14.8 | 2341 | 332 | 151 | 31 |
| 8 | 16.6 | 1590 | 225 | 128 | 18 |
| 9 | 17.1 | 2634 | 359 | 177 | 29 |
| 10 | 16.4 | 1404 | 189 | 113 | 16 |
| 11 | 16.2 | 2464 | 354 | 170 | 28 |
| 12 | 15.8 | 1971 | 272 | 160 | 26 |
| 13 | 18.8 | 1345 | 191 | 115 | 8 |
| 14 | 16.6 | 2454 | 326 | 166 | 27 |
| 15 | 13.6 | 2585 | 375 | 184 | 42 |
| 16 | 15.7 | 2769 | 380 | 198 | 33 |
| 17 | 14.1 | 2732 | 352 | 196 | 48 |
| 18 | 16.2 | 2765 | 399 | 198 | 35 |
| 19 | 13.5 | 2924 | 373 | 217 | 50 |
| 20 | 15.3 | 2587 | 348 | 192 | 40 |
| AVE | 15.59 | 2407.1 | 330 | 173 | 31.5 |
| SD | 1.59 | 551.21 | 75.85 | 32.67 | 10.81 |
| min | 12.5 | 1345 | 189 | 113 | 8 |
| max | 18.8 | 3523 | 481 | 233 | 50 |
| IQR | 1.9 | 564.5 | 63.25 | 40 | 11.75 |

## References

1- Perrotta, G. (2020). Psychotic spectrum disorders: definitions, classifications, neural correlates and clinical profiles. Annals of Psychiatry and Treatment, 4(1), 070–084.

2- Spallek, S., Birrell, L., Kershaw, S., Devine, E. K., & Thornton, L. (2023). Can we use ChatGPT for Mental Health and Substance Use Education? Examining Its Quality and Potential Harms. JMIR Medical Education, 9(1), e51243.

3- Ayers, J. W., Poliak, A., Dredze, M., Leas, E. C., Zhu, Z., Kelley, J. B., … & Smith, D. M. (2023). Comparing physician and artificial intelligence chatbot responses to patient questions posted to a public social media forum. JAMA internal medicine, 183(6), 589–596.

4- Lundin, R. M., Berk, M., & Østergaard, S. D. (2023). ChatGPT on ECT: Can Large Language Models Support Psychoeducation?. The journal of ECT, 39(3), 130–133.

5- Lee, E. E., Torous, J., De Choudhury, M., Depp, C. A., Graham, S. A., Kim, H. C., … & Jeste, D. V. (2021). Artificial intelligence for mental health care: clinical applications, barriers, facilitators, and artificial wisdom. Biological Psychiatry: Cognitive Neuroscience and Neuroimaging, 6(9), 856–864.

6- Fakhoury M. (2019). Artificial Intelligence in Psychiatry. Advances in experimental medicine and biology, 1192, 119–125. 10.1007/978-

7- Singhal, K., Tu, T., Gottweis, J., Sayres, R., Wulczyn, E., Hou, L., Clark, K., Pfohl, S.R., Cole-Lewis, H.J., Neal, D., Schaekermann, M., Wang, A., Amin, M., Lachgar, S., Mansfield, P.A., Prakash, S., Green, B., Dominowska, E., Arcas, B.A., Tomašev, N., Liu, Y., Wong, R.C., Semturs, C., Mahdavi, S.S., Barral, J.K., Webster, D.R., Corrado, G.S., Matias, Y., Azizi, S., Karthikesalingam, A., & Natarajan, V. (2023). Towards Expert-Level Medical Question Answering with Large Language Models. ArXiv, abs/2305.09617.

